# The external validity of four risk scores predicting 30-day mortality after surgery

**DOI:** 10.1101/2022.03.15.22272450

**Authors:** Frederick Torlot, Chang-Yang Yew, Jennifer R. Reilly, Michael Phillips, Dieter G. Weber, Tomas B. Corcoran, Kwok M. Ho, Andrew J. Toner

**Affiliations:** Royal Perth Hospital, Perth, Australia; Department of Anaesthesiology and Perioperative Medicine, Alfred Hospital, Melbourne, Australia; Department of Anaesthesia and Perioperative Medicine, Monash University, Melbourne, Australia; University of Western Australia, Perth, Australia

## Abstract

**Background:** Surgical risk prediction tools can facilitate shared-decision-making and efficient allocation of perioperative resources. Such tools should be externally validated in target populations prior to implementation.

**Methods:** Predicted risk of 30-day mortality was retrospectively derived for surgical patients at Royal Perth Hospital from 2014 to 2021 using the Surgical Outcome Risk Tool (SORT) and the related NZRISK (n=44,031, 53,395 operations). In a sub-population (n=31,153), the Physiology and Operative Severity Score for the enumeration of Mortality (POSSUM) and the Portsmouth variant of this (P-POSSUM) were matched from the Copeland Risk Adjusted Barometer (C2-Ai, Cambridge, UK). The primary outcome was risk score discrimination of 30-day mortality as evaluated by area-under-receiver operator characteristic curve (AUROC) statistics. Calibration plots and outcomes according to risk decile and time were also explored.

**Results:** All four risk scores showed high discrimination (AUROC) for 30-day mortality (SORT=0.922, NZRISK=0.909, P-POSSUM=0.893; POSSUM=0.881) but consistently over-predicted risk. SORT exhibited the best discrimination and calibration. Thresholds to denote the highest and second-highest deciles of SORT risk (>3.92% and 1.52-3.92%) captured the majority of deaths (76% and 13% respectively) and hospital-acquired-complications. Year-on-year SORT calibration performance drifted towards over-prediction, reflecting a decrease in 30-day mortality over time despite an increase in the surgical population risk.

**Conclusions:** SORT was the best performing risk score in predicting 30-day mortality after surgery. Categorising patients based on SORT into low, medium (80-90^th^ percentile) and high-risk (90-100^th^ percentile) can guide future allocation of perioperative resources. No tools were sufficiently calibrated to support shared-decision-making based on absolute predictions of risk.

## Introduction

Surgical risk prediction tools can facilitate shared-decision-making and efficient allocation of perioperative resources^1^. Many such tools have demonstrated excellent predictive performance in the populations they are developed in (internal validation) but perform less well in external validation studies without recalibration^2, 3^. For shared-decision-making, where a patient may decline an operation or treatment based on the quoted risk, accurate predictive performance across the entire risk range (calibration) is important^4^.

The advent of locally developed or commercially available tools to calculate risk scores and record surgical outcomes in large data sets allows performance evaluation (external validation) at a hospital level prior to implementation. This evaluation can guide which, if any, available risk tools can be successfully applied to the local population, or whether bespoke risk tools should be developed. Furthermore, local data can define appropriate risk thresholds, above which patients can systematically be allocated extra resources with the aim of improving outcomes.

At Royal Perth Hospital, a data and digital innovation unit was recently established, linking a range of perioperative information in a single data warehouse. In parallel, the Copeland Risk Adjusted Barometer (C2-Ai, Cambridge, UK) system was introduced, primarily to evaluate risk-adjusted surgical outcomes and benchmark them against other hospitals in the system database. We set out to use these information systems to externally validate the performance of four common surgical risk tools, when applied in our hospital over a 7-year period.

## Methods

The study protocol, incorporating a waiver of consent, was approved by the Royal Perth Hospital Human Research Ethics Committee on the 12^th^ August 2021 (RGS0000004853). This retrospective observational study is reported in accordance with the Transparent Reporting of a Multivariable Prediction Model for Individual Prognosis or Diagnosis (TRIPOD) statement^5^.

### Risk tools assessed

Four commonly used risk scores were evaluated for the prediction of 30-day mortality. First, version one of the Surgical Outcome Risk Tool (SORT)^6^ and the related NZRISK tool^2^ were evaluated, as they were recently identified as suitable candidates for adaptation in Australian hospitals^7^ and their component variables were available within the data and digital innovation warehouse. Second, the Physiology and Operative Severity Score for the enumeration of Mortality (POSSUM)^8^ and the Portsmouth variant of this (P-POSSUM)^9^ were evaluated as they were available within the Copeland Risk Adjusted Barometer system. This system applies proprietary algorithms to derive risk scores retrospectively from coding-based information after hospital discharge.

### Eligibility, data extraction and selection

Surgical episodes taking place between July 1^st^ 2014 and June 30^th^ 2021 were examined if the following eligibility criteria were met: age ≥18 years; procedure undertaken in the main theatre complex by a surgeon; planned postoperative stay ≥1 night; non-indigenous ethnicity (to allow a fair comparison across risk scores with varying approaches to ethnicity).

The perioperative data required to apply the SORT and NZRISK tools were extracted from the data and digital innovation warehouse. Sources of data in this extract included the Theatre Management System and WebPas software used across public hospitals in Perth to schedule and track operations and outcomes. These were supplemented by the International Classification of Diseases 10^th^ Revision Australian Modification (ICD-10-AM) codes^10^ and the Australian Classification of Health Interventions (ACHI) procedure codes^11^, entered by the coding department after the end of an inpatient episode of care.

The data extract was further refined by taking the following sequential steps: Step 1, removal of all operative data other than the first surgery within an episode of care; Step 2, removal of procedures performed under local anaesthesia or sedation without an upper or lower limb block; Step 3, removal of surgical specialties that were not included in the original risk score development studies or were not fully represented during the study period; Step 4, removal of procedures with missing SORT or NZRISK variables; Step 5, matching of procedures captured by the Copeland Risk Adjusted Barometer; Step 6, restriction to the first episode of surgical care within the 7-year study period.

### Calculating 30-day mortality risk

The SORT and NZRISK predicted risks of 30-day mortality were calculated by mapping the component variables for each eligible surgical episode from the data warehouse extract (Supplementary Table 1). Age, sex and surgical urgency were obtained directly from fields in the Theatre Management System. Other variables required processing of coded data. Specifically, ACHI procedure codes were matched to a John Hopkins Pasternak Operative Severity Score (1-5)^12^ applying the classification table prepared and used by the NZRISK group^2^. If multiple procedures were coded for the same operative episode, the procedure with the highest severity score was selected to represent the overall complexity. The ACHI procedure code block denoting the body or organ system targeted by that procedure was used to assign the surgical speciality type, again following the NZRISK methodology. The ACHI anaesthetic code provided the American Society of Anesthesiologists (ASA) physical status score, but if this was missing, the value entered in the Theatre Management System during the operation was used. Finally, ICD-10-AM codes recorded during care at Royal Perth Hospital in the five years preceding an eligible surgical episode were examined, and if codes were present in the C01-C96 or D00-D09 range, then the patient was determined to have a positive cancer status.

After variable mapping, the open-access regression equations outlined below were used to summate the risk score and calculate the 30-day mortality prediction for each patient.

SORT predicted risk of 30-day mortality (R): ln (*R*/(1−*R*)) = -7.336 + risk score

NZRISK predicted risk of 30-day mortality (R): ln (*R*/(1−*R*)) = -10.625 + risk score

### Outcome Data

Routinely collected outcome data was extracted from the data and digital innovation warehouse. This included 30-day mortality, hospital acquired complications (HACs), intensive care unit (ICU) bed hours and length of stay. As per Australian government policy (www.safetyandquality.gov.au/our-work/indicators/hospital-acquired-complications), HACs are recorded if the required ICD-10-AM code(s) and the condition onset are linked to an episode of care. As the criteria to define a HAC evolves over time, the most recent iteration at the time of the analysis (Excel Groupers – Version 3.0) was applied across the whole data set. Days alive and out of hospital in the first 30 days (DAH-30), estimated at Royal Perth Hospital from public healthcare facility admission data in the metropolitan area, was also extracted.

### Statistical Analysis

Predictive performance was evaluated with receiver operator characteristic curves and calibration plots, restricted to the first episode of care in the study period if multiple episodes took place (avoiding violation of the independent and identically distributed data assumption). The primary outcome was risk score discrimination of 30-day mortality as evaluated by area-under-receiver operator characteristic curve (AUROC) statistics. Calibration plots were constructed to examine predicted risks against observed risks. Non-parametric smoothed best-fit curves were added to calibration plots to aid visual evaluation. Calibration plots were re-scaled to reflect the risk range where predictions were relatively precise. To assess predictive performance year-on-year, select risk score(s) were further evaluated with calibration plots using annualised data.

Surgical population risk and 30-day mortality trends over time were assessed with a mixed-effects regression model, applied to all episodes of care within the 7-year study period. Surgical outcomes in the highest and next-highest deciles of risk were also compared to lower risk patients, applying mixed-effects models with either logistic or negative binomial regression as appropriate.

Analyses were completed in Stata (StataCorp. 2019. *Stata Statistical Software: Release 16*. College Station, TX: StataCorp LLC).

## Results

The number of surgical episodes meeting eligibility criteria for the data extract was 70,846 (Figure 1). After the various exclusion criteria were applied, SORT and NZRISK predicted 30-day mortality were calculated for 44,031 patients, undergoing 53,395 distinct surgical episodes. Mortality according to patient and operative characteristics in the total population is summarised in Table 1.

**Figure 1.**
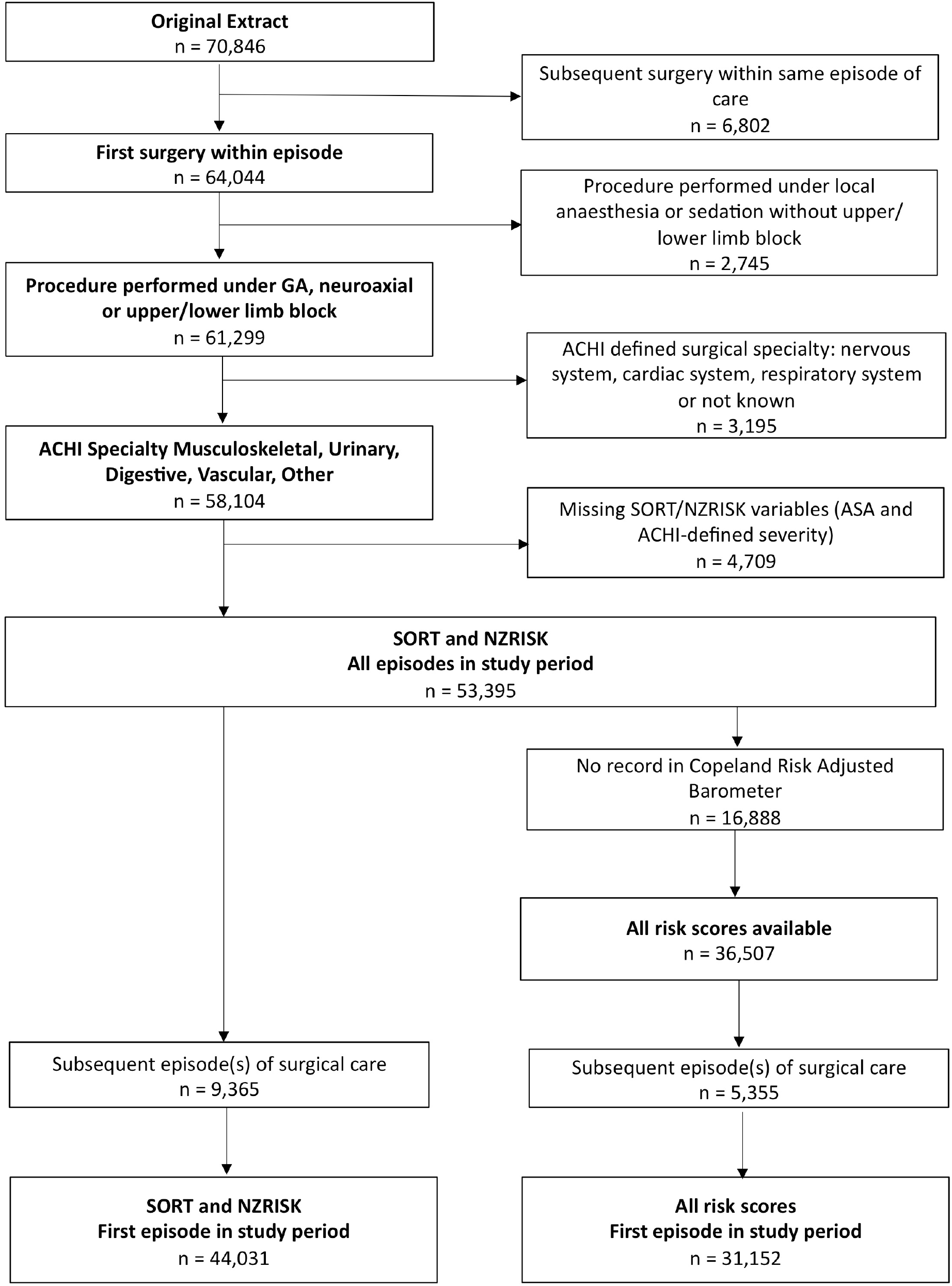
Flow diagram for data extraction and selection. GA, General Anaesthesia; ACHI, Australian Classification of Health Interventions; ASA, American Society of Anesthesiologists physical status score.

**Table 1.** Patient and operative characteristics in the total study population

|  | Surgical Episodes, n=53,395 | Deaths, n (%) |
| --- | --- | --- |
| Age (years) |  |  |
| <65 | 36210 | 97 (0.27) |
| 65-79 | 11745 | 153 (1.30) |
| ≥80 | 5440 | 308 (5.66) |
| Sex |  |  |
| Male | 31664 | 332 (1.05) |
| Female | 21731 | 226 (1.04) |
| ASA physical status |  |  |
| 1 | 11020 | 3 (0.03) |
| 2 | 24196 | 25 (0.10) |
| 3 | 15719 | 239 (1.52) |
| 4 | 2413 | 270 (11.19) |
| 5 | 47 | 47 (44.68) |
| Surgical severity |  |  |
| Minor/Intermediate/Major | 46917 | 397 (0.85) |
| Xmajor/complex | 6478 | 161 (2.49) |
| Surgical urgency |  |  |
| Elective | 19846 | 68 (0.34) |
| Expedited | 23820 | 300 (1.26) |
| Urgent | 8993 | 156 (1.73) |
| Immediate | 736 | 736 (4.62) |
| Surgical type |  |  |
| Other | 12396 | 33 (0.27) |
| Musculoskeletal | 23905 | 317 (1.33) |
| Urological | 5086 | 32 (0.63) |
| Digestive | 10166 | 137 (1.35) |
| Vascular | 1842 | 39 (2.12) |
| Cancer in last 5 years |  |  |
| No | 44986 | 451 (1.00) |
| Yes | 8409 | 107 (1.27) |

Matching to the Copeland Risk Adjusted Barometer resulted in 31,152 patients with all four risk scores available. In this population, the risk scores showed high discrimination (AUROC) for 30-day mortality (SORT=0.922, NZRISK=0.909, P-POSSUM=0.893; POSSUM=0.881) but consistently over-predicted risk (Figure 2). On visual inspection of calibration plots in the 0-10% range, SORT was marginally better calibrated than NZRISK and substantially better calibrated than P-POSSUM OR POSSUM. The proportion of patients with predicted 30-day mortality >10% was low for all the risk scores (SORT=2.87%, NZRISK=3.70%, P-POSSUM = 4.98% and POSSUM = 5.98% respectively), resulting in widened confidence intervals in the 10-100% range (Supplementary Figure 1).

**Figure 2.**
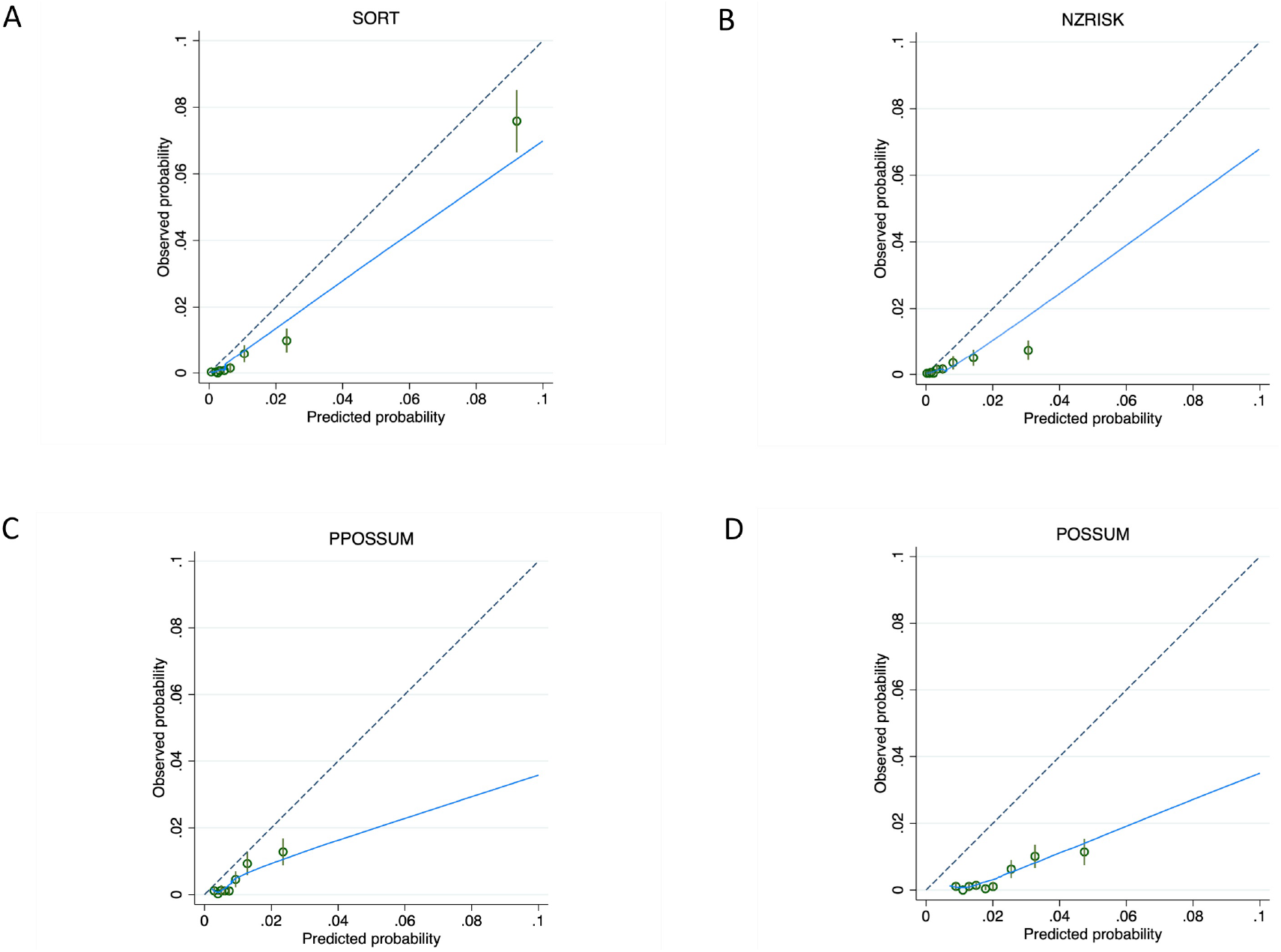
Calibration plots for predictions of 30-day mortality between 0-10% using SORT (A), NZRISK (B), P-POSSUM (C) and POSSUM (D). Dashed line represents perfect calibration, blue line is non-parametric smoothed best-fit curve, green bars are 95% confidence intervals.

As SORT exhibited the best discrimination and overall calibration, and was also the most parsimonious risk score, further in-depth analyses focused on this measure only. Annualised evaluation of SORT revealed good visual calibration in the 0-10% risk range in the first three years of the study period, followed by a steady calibration drift towards over-predicted risk (Figure 3). This drift reflected a significant decrease in annualised 30-day mortality over time (OR 0.865 per elapsed year, 95% CI 0.828-0.903, P<0.001) despite an increase in the predicted 30-day mortality (0.073% increase per elapsed year, 95% CI 0.057-0.089%, P<0.001) - see Figure 4.

**Figure 3.**
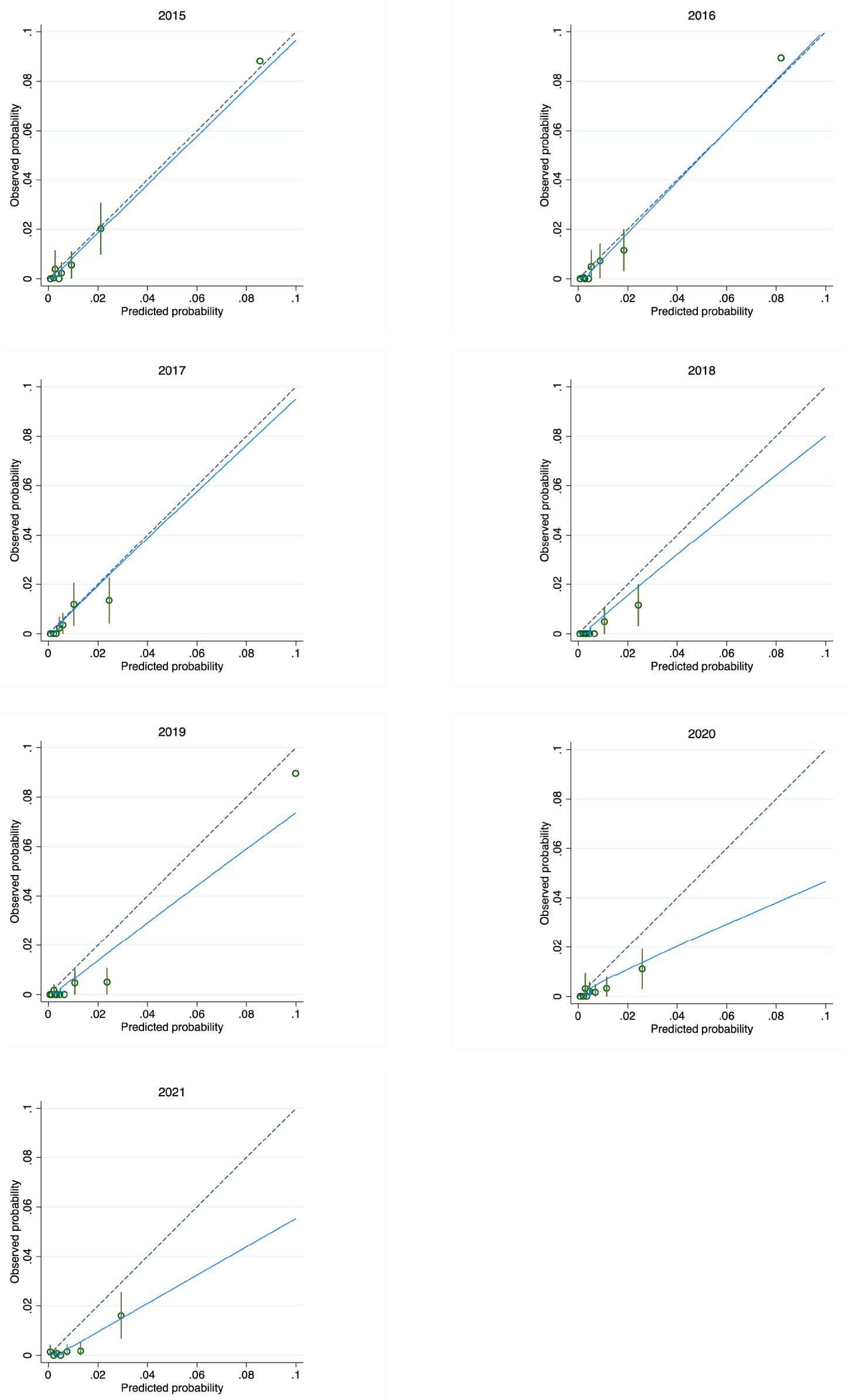
Annualised calibration plots for SORT predictions of 30-day mortality according to financial year end (1^st^ July to 30^th^ June). Dashed line represents perfect calibration, blue line is non-parametric smoothed best-fit curve, green bars are 95% confidence intervals.

**Figure 4.**
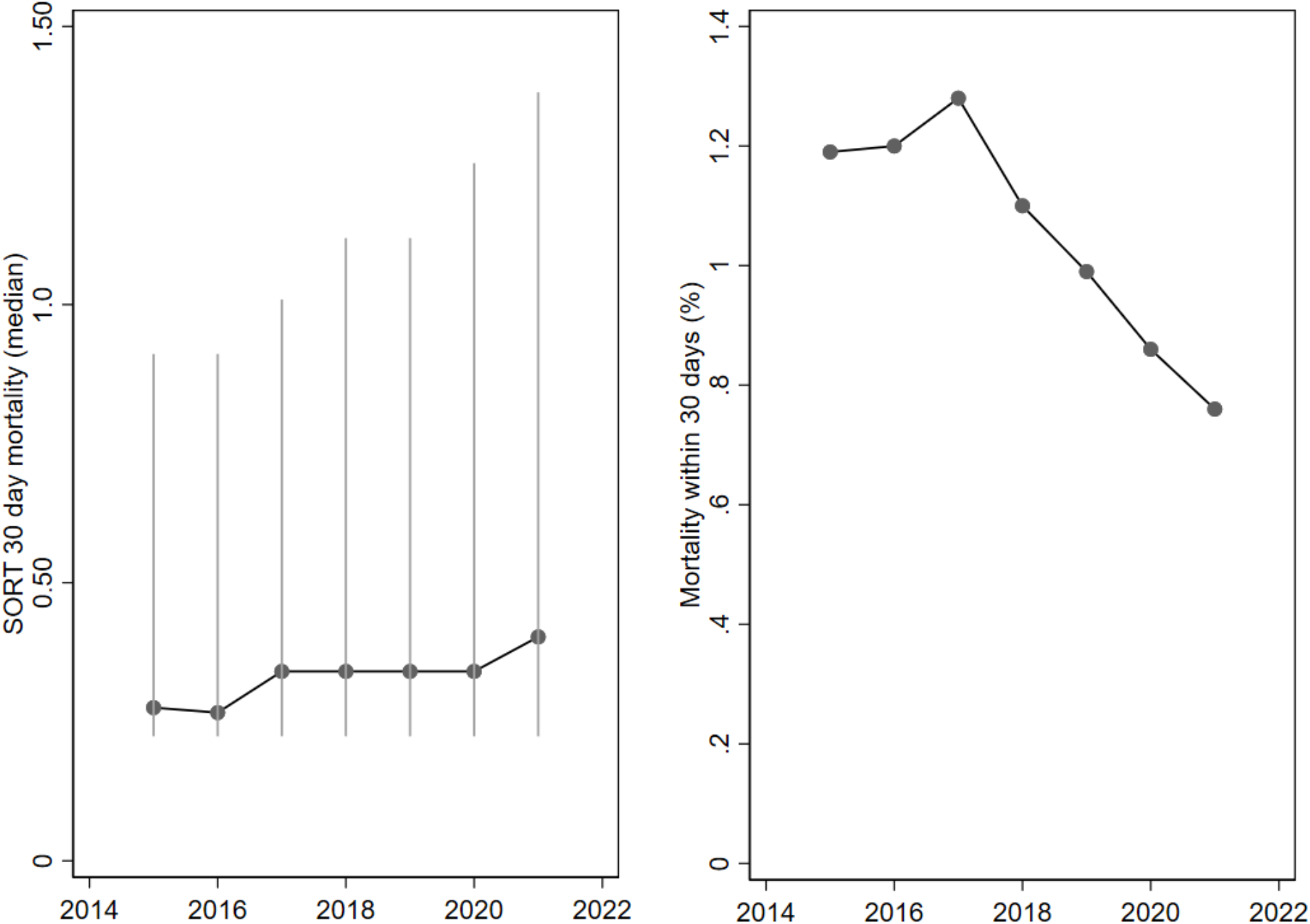
Annualised median (IQR) SORT risk and 30-day mortality

Thresholds to denote the highest and next-highest deciles of risk (SORT 30-day mortality predicted > 3.92% and 1.52-3.92%) across the 7-year period captured the majority of deaths (76% and 13% respectively). Applying these thresholds to classify low, medium and high-risk surgical patients resulted in a very significant increase in the odds of 30-day mortality and major hospital acquired complications in the medium and high risk categories, manifesting in prolonged length of hospitalisation and less days alive and out of hospital (Table 2). The likelihood of hospital acquired complications indicative of a greater nursing burden also increased across the risk groups (Supplementary Table 2).

**Table 2.** Mortality and major morbidity in low, medium and high SORT risk groups

|  | Low Risk | Medium Risk | High Risk |
| --- | --- | --- | --- |
|  | SORT <1.52% | SORT 1.52-3.92% | SORT >3.92% |
|  | n=42,705 | n=5,516 | n=5,174 |
| Mean predicted 30-day mortality (%) | 0.394 | 2.46 | 9.84 |
| Median predicted 30-day mortality (%) | 0.266 | 2.38 | 6.63 |
| Observed 30-Day Mortality, n (%) | 59 (0.14) | 74 (1.34) | 425 (8.21) |
| Odds ratio (95% CI) |  | 11.1 (7.45-16.6) | 98.2 (54.7-176) |
| Any Hospital Acquired Complication, n (%) | 1203 (2.82) | 694 (12.6) | 1110 (21.5) |
| Odds ratio (95% CI) |  | 5.20 (4.67-5.79) | 10.3 (9.23-11.5) |
| Healthcare associated infection, n (%) | 635 (1.49) | 384 (6.96) | 519 (10.0) |
| Odds ratio (95% CI) |  | 5.20 (4.52-5.99) | 7.98 (6.96-9.16) |
| Delirium, n (%) | 200 (0.47) | 156 (2.83) | 330 (6.38) |
| Odds ratio (95% CI) |  | 6.31 (5.07-7.85) | 15.3 (12.5-18.7) |
| Cardiac Complications, n (%) | 141 (0.33) | 120 (2.18) | 218 (4.21) |
| Odds ratio (95% CI) |  | 6.71 (5.25-8.58) | 13.3 (10.7-16.4) |
| Respiratory Complications, n (%) | 144 (0.34) | 112 (2.03) | 202 (3.90) |
| Odds ratio (95% CI) |  | 6.27 (4.84-8.12) | 12.6 (9.84-16.1) |
| Venous Thromboembolism, n (%) | 88 (0.21) | 36 (0.65) | 45 (0.87) |
| Odds ratio (95% CI) |  | 3.18 (2.16-4.69) | 4.25 (2.96-6.09) |
| Gastrointestinal Bleeding, n (%) | 27 (0.06) | 21 (0.38) | 45 (0.87) |
| Odds ratio (95% CI) |  | 6.04 (3.41-10.7) | 13.9 (8.59-22.4) |
| Surgical complications requiring unplanned return to theatre, n (%) | 76 (0.18) | 36 (0.65) | 44 (0.85) |
| Odds ratio (95% CI) |  | 3.74 (2.48-5.64) | 4.98 (3.37-7.35) |
| Renal Failure | 4 (0.01) | 8 (0.15) | 18 (0.35) |
|  |  | 15.5 (4.67-51.4) | 37.2 (12.6-110) |
| ICU admission, n (%) | 1339 (3.14) | 751 (13.6) | 1023 (19.8) |
| Odds ratio (95% CI) |  | 5.46 (4.86-6.13) | 9.17 (8.11-10.4) |
| Length of hospital stay in days, Median (IQR) [Range] | 2.0 (1.2-4.2) [0.1-283.6] | 5.1 (2.3-9.2) [0.2-300.0] | 6.6 (3.9-12.1) [0.1-211.1] |
| Coefficient, (95% CI) |  | 0.723 (0.693-0.754) | 1.01 (0.986-1.04) |
| DAH-30, Median (IQR) [Range] | 29 (26-29) [0-30] | 25 (18-28) [0-30] | 20 (6-25) [0-30] |
| Coefficient, (95% CI) |  | -5.00 (-5.25- -4.75) | -10.0 (-10.3- -9.75) |

All odds ratios and 95% confidence intervals (CI) reflect a pair-wise comparison with the low risk group and have a significance level of $P < 0.001$ .

## Discussion

In this large single-centre study, SORT, NZRISK, P-POSSUM and POSSUM risk scores exhibited high levels of discrimination for 30-day mortality for patients undergoing surgery, but calibration displayed varying degrees of over-prediction. The SORT mortality risk demonstrated the best external validity in our population and proved an effective basis for a broad categorisation of patients into low, medium and high-risk, based on predicted risk deciles. These categories were associated with hospital acquired complications, length of stay and derived DAH-30, in addition to 30-day mortality. Whilst it has long been recognised that the majority of postoperative complications and deaths occur in a minority of high-risk patients^13^, there is a paucity of data in the literature on how thresholds for a specific risk score should be set to identify such patients. Indeed, as hospitals will vary in the number of high-risk surgical patients they encounter annually and the resources available to them, thresholds to designate high-risk can only be usefully set at an institution level in order to identify a manageable patient volume that can reliably receive enhanced care. This study demonstrates how the advent of large hospital-level risk and outcome data sets facilitates such an approach.

The calibration findings in our study are consistent with recent prospective studies that report suboptimal risk score calibration in surgical populations beyond the original development populations^2, 3^. It is only when risk equations are recalibrated or refined within a target population that subsequent risk scores meet the exacting calibration standards required to support shared-decision-making on the basis of absolute risk. Despite the suboptimal calibration reported in this and other external validation studies, it can be argued that the levels of predictive performance observed are sufficient to support shared-decision-making on the basis of *relative* risk, either by classifying patients into broad risk groups, or by interpreting absolute risk predictions alongside contemporaneous calibration plots. In this regard, given the low number of patients with predicted 30-day mortality above 10%, calibration plots to assist shared-decision-making should incorporate confidence intervals at each level of predicted risk or be restricted to the 0-10% risk range.

In contrast to the calibration observed using the entire data set, an annual analysis of SORT demonstrated excellent calibration for the first three years of the study period (1^st^ July 2014 to 30^th^ June 2017). It is worth noting that the original version of SORT was derived from 2010 data in the UK and first published in 2014. After 2017, a steady calibration drift towards over-predicted risk was observed. This drift reflected a significant decrease in annualised 30-day mortality despite an increase in the surgical population risk. Similar trends should be evident in most high-performing hospitals, where the patients treated become older and more co-morbid with time, but perioperative processes improve. For example, at Royal Perth Hospital a number of initiatives were implemented and completed during the study period, including a “Safety Afterhours For Everyone” (SAFE) team to enhance management of patients at risk of clinical deterioration. The observed calibration drift highlights the importance of incorporating periodic recalibration into systems aiming to maximally support shared-decision-making.

The acquisition of institution-level risk and event rate data as in our study can guide strategic decisions with respect to the allocation of perioperative resources. For example, one of the most common clinical applications of surgical risk prediction is to decide who is admitted to an ICU bed on the day of surgery. This type of intervention is expensive (approximately A$5,000 per bed-day^14^) and often captures only a small portion of the at-risk period before ward discharge^15^. Even optimistically assuming a 25% relative risk reduction in patients experiencing at least one hospital acquired complication by allocation of a postoperative ICU bed, implementing this intervention in all high-risk patients at Royal Perth Hospital where the current default is a standard postoperative ward bed, requires $3.7 million dollars or $99,000 per complication prevented. Although formal evaluations would also include the cost-savings from preventing hospital acquired complications, it is clear that more cost-effective interventions that span the entire at-risk period and accommodate increased nursing requirements are needed. One such intervention introduced in 2021 in our hospital is the availability of enhanced postoperative ward beds that incorporate remote monitoring of vital signs and automated clinical deterioration alerts. Future studies are planned to evaluate the impact of this Healthcare in a Virtual Environment (HIVE) approach.

A limitation of this study is the retrospective calculation of the risk scores, derived in large part from hospital administrative data after discharge from the index hospital episode. In particular, we were only able to obtain a cancer diagnosis in the last five years if the patient had been treated as an inpatient at Royal Perth Hospital and if this diagnosis had been coded. A more reliable indicator of this cancer field might in future be obtained via the state-wide data linkage system. It is therefore possible that the predictive performance limitations we observed, in particular the calibration findings, reflect inaccuracies in the retrospective methodology and that prospectively acquired risk scores would perform better. However, in their recent prospective study Wong and colleagues reported remarkably similar findings to the current work, showing that SORT outperformed other more complex risk scores for 30-day mortality including P-POSSUM, with all risk scores over-predicting risk. These similarities indicate that surgical risk depends consistently on a small number of objective variables that do not change perioperatively^16^, and coding approaches to risk estimation that can detect such objective variables are likely valid. As most hospitals either currently collect such data, or will with increasing healthcare digitisation, we consider integrated national and institution-specific risk prediction systems highly feasible.

Some of the retrospective outcome data collected in our study also has limitations. The nationally determined methodology to record hospital acquired complications will underestimate morbidity relative to prospective audit^17^. This is especially true where clinical note keeping is lacking or where complications that occur frequently after hospital discharge are assessed, such as surgical site infection^18^. Furthermore, our measure of DAH-30 doesn’t account for admissions in the private, non-metropolitan sectors, and likely over-estimates performance. Nevertheless, these methodological limitations are consistent for all our patients and thus provide useful insights into the outcome differences across surgical risk groups.

In conclusion, SORT, NZRISK, P-POSSUM and POSSUM risk scores exhibited high levels of discrimination but suboptimal calibration for 30-day mortality at Royal Perth Hospital over a 7-year period. SORT was the best performing surgical risk tool and effectively categorised patients into low (0-80^th^ percentile), medium (80-90^th^ percentile) and high (90-100^th^ percentile) risk. Defining these risk category thresholds for efficient and reliable allocation of perioperative resources is the key advantage of locally developed surgical risk and outcome databases. Risk tools sufficiently calibrated for shared-decision-making based on absolute risk may also be feasible but will likely require the development of region or institution specific risk models that incorporate periodic recalibration.

**Manuscript under review, BJA Open**

## Data Availability

All data produced in the present work are contained in the manuscript

## Acknowledgements

<u>Access to NZRISK surgical severity classification table</u>

D Campbell, Auckland City Hospital, Auckland

L Boyle, University of Auckland, Auckland

<u>Extracts from data and digital innovation warehouse</u>

J Mamas, Royal Perth Hospital, Perth

G North, Royal Perth Hospital, Perth

<u>Development of DAH-30 at Royal Perth Hospital</u>

D Brooke, Royal Perth Hospital, Perth

## Author’s contributions

FT: Data extraction, data processing and revision of paper

CYY: Data processing, statistical analysis and revision of paper

JR: Data interpretation and revision of paper

MP: Statistical analysis

DW: Data interpretation and revision of paper

TC: Data interpretation and revision of paper

KH: Data interpretation and revision of paper

AT: Study conception and design, data extraction and processing, first draft of paper

---

**Supplementary Figure 1.**
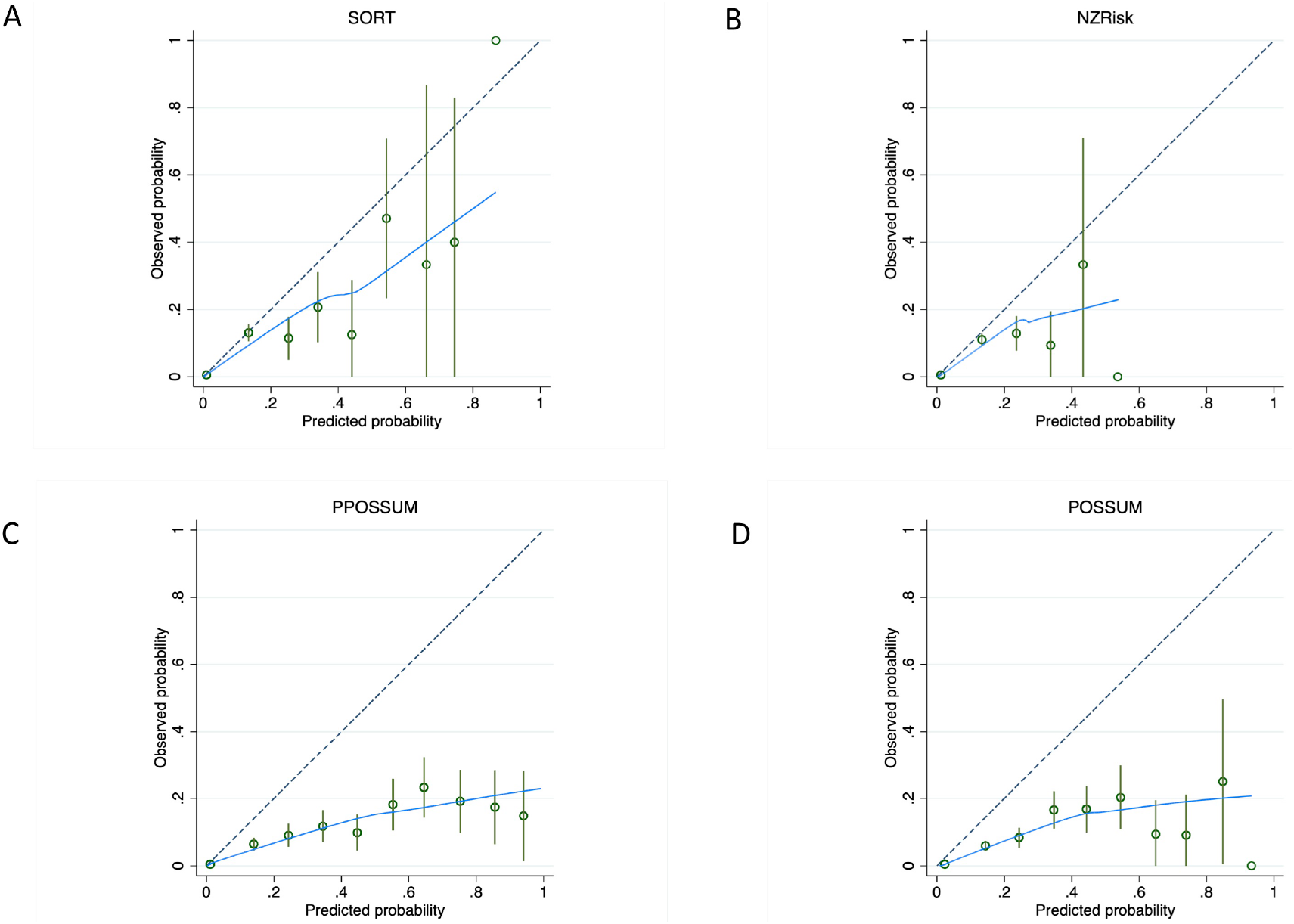
Calibration plots for predictions of 30-day mortality between 0-100% using SORT (A), NZRISK (B), P-POSSUM (C) and POSSUM (D). Dashed line represents perfect calibration, blue line is non-parametric smoothed best-fit curve, green bars are 95% confidence intervals.

**Supplementary Table 1.** Mapping raw data from information storage systems to SORT and NZRISK scores

| Raw data | Processed data | SORT Risk Score | NZRISK Risk Score |
| --- | --- | --- | --- |
| ACHI Procedure Code(s) | Transformed to “Surgical grade 1-5” | Surgical grade 4/5 =<br>Xmajor/complex =<br>0.381 | Surgical grade 4/5<br>= 0.592 |
| ACHI Procedure Code(s) | Transformed to NZRISK<br>“Surgical Specialty” | Digestive or Vascular<br>= Thoracics,<br>gastrointestinal or<br>vascular surgery =<br>0.712 | Musculoskeletal =<br>0.319<br>Urinary = 0.391<br>Digestive = 0.640<br>Vascular = 0.966 |
| ACHI Anaesthetic ASA<br>Code(S)/ TMS ASA Entry | Transformed to “Merged<br>ASA 1-5” | Grade 3 = 1.411<br>Grade 4 = 2.388<br>Grade 5 = 4.081 | Grade 3 = 0.448<br>Grade 4/5 = 0.752 |
| TMS Operation Urgency | Transformed to “SORT<br>Urgency”<br>Elective = Elective<br>Expedited = Urgent or<br>Emergency<48h<br>Urgent = Emergency or<br>Emergency <24h or <6h<br>Immediate = Emergency<br><2h or <15min | Expedited = 1.236<br>Urgent = 1.657<br>Immediate = 2.452 | Expedited or<br>Urgent or<br>Immediate =<br>Emergency = 2.236 |
| ICD-10 Codes for last 5<br>years | Transformed to “Cancer” | Cancer = 0.667 | Cancer = 0.936 |
| TMS Age at operation | N/A | 65-79 years = 0.777<br>≥80 years = 1.591 | Age = 0.060 per<br>year |
| TMS Patient Gender | N/A | N/A | Male sex = 0.018 |

ACHI, Australian Classification of Health Interventions; TMS, Theatre Management System; ICD-10, International Classification of Diseases 10<sup>th</sup> Revision

**Supplementary Table 2.**
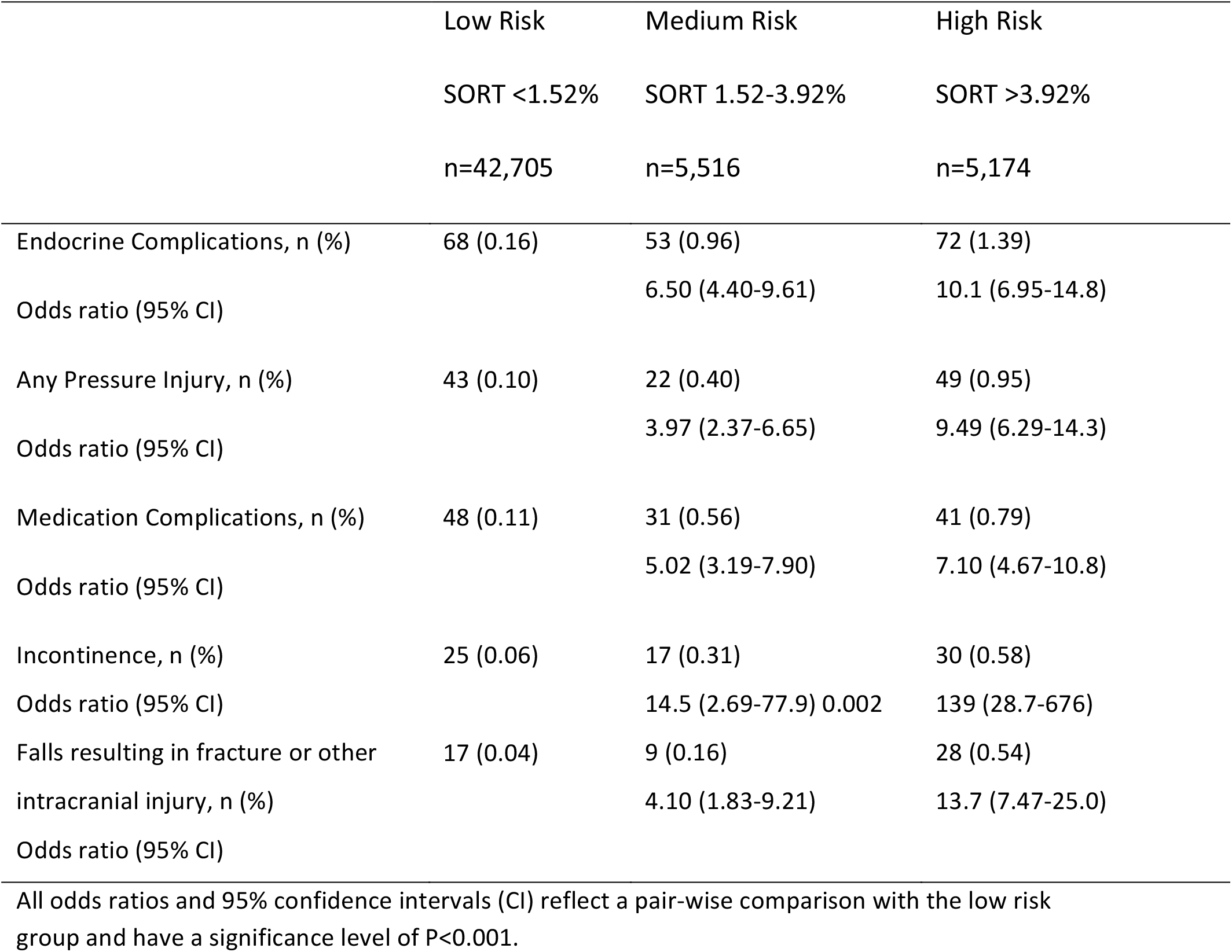
Hospital acquired complications linked to a high nursing burden in low, medium and high SORT risk groups

